# Perceptions and Outcomes of a Hospital Medicine (HM) Advanced Practice Provider (APP)-Led Care Model: A Qualitative Study

**DOI:** 10.64898/2026.02.18.26346538

**Authors:** Alisha T. DeTroye, Elizabeth Tysinger, Jacqueline Lippert, Kelly Conner, Chris Gillette

## Abstract

**Background:** A Hospital Medicine Advanced Practice Provider (HMAPP)-led care model developed in response to the high acuity and increased patient volumes associated with the Covid-19 pandemic. Although anecdotally perceived as a successful model, questions remained if there was adequate pre-planning and formal implementation strategy for stakeholder buy-in.

**Objective:** To elicit HM physicians’ and APPs’ perceptions of the HMAPP-led care model implementation and consider necessary steps for optimal future APP care model development and operation.

**Design, Setting and Participants:** This qualitative study used 10 (5 physicians and 5 APPs involved in the care model pre- and post-implementation) individual semi-structured, virtual interviews based on the Consolidated Framework for Implementation Research (CFIR). Deductive and inductive rapid analysis was utilized to analyze the data.

**Results:** Two themes emerged as strengths: 1) Experienced APPs delivered the care model, 2) Acceptance of the care model evolved over time.

Four themes suggested opportunities for future development: 1) Guidelines should expand from patient distribution to include minimal collaboration and escalation expectations, 2) Culture change was a barrier to model implementation and acceptance, 3) Intentional collaboration between APPs and Physicians is necessary, 4) Investment in standardized onboarding enhances buy-in of the care model.

**Conclusion:** The impact of an APP care model can be elevated if implemented with key principles and strategies. This is critical in an evolving health care landscape where all providers need to collaborate and practice with their full expertise to maximize safe, efficient and quality patient care.

## INTRODUCTION

Hospital Medicine (HM) has only been in existence for approximately 30 years, making it one of the newest medical subspecialties.^1^ Originally the goal of HM was to provide a more specialized level of care as well as continuity of care as patients’ needs became more complex.^2^ HM teams have expanded to include Advanced Practice Providers (APPs) due to increasing patient cases and resident hour restrictions to ensure quality patient care, to address length of stay and rising costs of care.^3,4^ In 2024, 3.5% (n=4,756) of all PAs nationally identify HM as their clinical specialty and 2.9% of NPs are certified in acute care, highlighting the importance of APPs at providing efficient, safe, and high quality inpatient care.^5, 6^

While systems have heavily invested in HMAPPs due to rising demand and a projected HM physician shortage, questions remain about how to best integrate HMAPPs into current inpatient workflows. The dramatic effects of the Covid-19 pandemic necessitated rapid changes in workflows affecting all inpatient providers. ^7, 8, 9, 10^

Many of these care models, however, were appropriately developed rapidly in response to a global pandemic but their effectiveness may be hindered by a lack of focus on how the new workflows impact patient care teams.^9^ Westergaard et al conducted focus groups with HM physicians, APPs and system leadership at multiple institutions to obtain information on how APPs are utilized on HM services and how new billing practices might impact physician-APP collaboration. The findings indicated that external drivers, such as staffing shortages, finances and policy influenced HMAPP deployment but failed to consider how these changes impacted team dynamics, such as physician and APP collaboration. Since HMAPP care models continue to evolve rapidly with heterogeneous implementation there is a need to further evaluate how changes in HM APP deployment impact team design.^7^ Prior work indicates APP-led care models have comparable patient outcomes (direct costs, 30-day readmissions, mortality) and perceived patient satisfaction to hospitalist physician or resident models.^11-13^

In APP role development, it is important to consider best practice for onboarding, initial and ongoing competency and professional development to enhance APP recruitment and retention.^8,14^ This can be achieved through formal transition to practice programs as well as informal mentoring. Additionally, creating guidelines for role clarity, team management, daily workflow and optimization of team collaboration and dynamics, the hospital medicine team can follow standard best practice of an APP care model. ^15^ Robust onboarding programs, well defined APP roles and responsibilities and opportunities for growth enhance APP job satisfaction and reduce turnover.^4^

Like other academic medical centers (AMCs), Atrium Health Wake Forest Baptist (AHWFB) created a new APP-led care model in 2020 to address increasing volumes and complexity of patients during the Covid-19 pandemic. Anecdotally, the model is perceived as a success, allowing APPs to expand patient access to care in the hospital setting, through more autonomous practice, like other reports in the literature.^16^ As stated above, current HM APP deployment is generally a result of external forces instead of planned, staged implementation using stakeholder engagement. Implementation Science (IS) is the study of methods to promote the adoption and use of evidence-based practices into routine patient care.^17^ Improving our understanding of how to integrate HM APPs into inpatient care using stakeholder engagement and iterative implementation has the potential to not only impact inpatient care team models but also improve patient access and care quality. The goal of this study, therefore, was to evaluate provider perceptions since the APP care model development in 2020 guided by the Consolidated Framework for Implementation Research (CFIR).^18^

## METHODS

### Study Design

The Wake Forest University School of Medicine Institutional Review Board (# 00122461) approved this study. The study team utilized the COREQ checklist to guide study reporting.^19^ We used an interpretivist paradigm to better understand health care providers’ perceptions about the use of an APP-led HM care model. We developed interview questions to explore perceptions of safety and effectiveness as well as provider experience with an APP-led AMC HM care model.

### Interview Guide Development

The CFIR framework served as the basis for development of the interview guide. This IS framework has been utilized to determine perceptions of the success or failure of implementation.^18^ Reviewing the literature, helped to frame the intersection of inner and outer domain influence within the CFIR constructs.^18, 20^ Prior to the actual interviews, a mock interview was conducted with one of the investigators (JL) who was involved with the HM APP care model development and implementation. Feedback from the mock interview helped to shape the verbiage and interview strategy. Since semi-structured interviews were conducted, additional related topics could be explored outside of the guide as directed by the interviewee. The full interview guide is provided in Appendix A.

### Setting and Participants

We conducted semi-structured interviews with physicians and HM APPs at one AMC using the Microsoft Teams platform, part of Microsoft 365. The study team purposely sampled providers to ensure providers had pre- and post-implementation experience. The eligible HM providers included physician assistants, nurse practitioners, and physicians, some of whom held department and system leadership roles. Providers who were not working on the HM service prior to care model implementation were not eligible to participate. Potential interviewees received an email from the study team explaining the study. To minimize variation in participant inclusion, the first author utilized a snowball technique. *A priori*, we anticipated needing to conduct 20 interviews to ensure thematic saturation; however, saturation was achieved with 10 interviews conducted, consistent with prior qualitative sample size literature.^21^

### Reflexivity

The research team consisted of a physician assistant and institutional APP leader, a nurse practitioner, HM physician, and PhD researcher. These unique backgrounds of the study team contributed to qualitative design, framed the research questions and invested in improving the process of IS of care model development. It was acknowledged and disclosed with the consent that the researchers may have prior professional interactions with participants. Each participant was assigned a unique deidentified code to maintain confidentiality and objectivity of analysis.

### Data Collection Procedure

A trained qualitative interviewer and health care provider (ET) conducted the interviews with participants. Prior to the start of each interview, the interviewer explained the interview process with each interviewee. Interviews were designed to last about 60 minutes each. Each interview was recorded and transcribed by the Microsoft Teams platform. During the interviews the primary investigator (AD) captured field notes off camera, while the nurse practitioner co-investigator (ET) conducted the interview face to face on Microsoft Teams.

### Data Analysis and Validation

To maximize efficiency but maintain a comprehensive approach, we used a rapid inductive and deductive analysis approach guided by CFIR.^20, 22^ The primary investigator (AD) coded the findings within 48 hours to capture interview themes and assigned scores for each domain and construct identified. Score ratings were based on impact, which is positive or negative influence on implementation and strength, which is weak or strong influence on implementation. Ratings ranged from +2 to -2 with 0 representing neutral or opportunity score. ^22^ The primary investigator(AD) captured a summary of the findings, including relevant quotes and rationale for score recorded. The secondary analyst, who was the interviewer (ET), reviewed the template and listened to the recordings to provide edits to the thematic analysis and clarify relevant quotes. The two researchers met weekly to review discrepancies and reach consensus. A third researcher (CG) was available for validation or “tie breaking.” Themes and specific quotes were sent to the participants for member checking to ensure perspectives were accurately captured and interpreted by the research team. Once all individual data was analyzed and scores assigned, the overall frequency of themes by aggregate scores was captured. Then the pooled data was analyzed to compare differences in themes between APP and Physician participants. Microsoft Excel, a part of Microsoft 365, was used to manage data.

## RESULTS

The potential study population was thirty providers (19 physicians, 9 PAs and 2 NPs) who practiced at the AMC HM service pre- and post-implementation. We recruited five physicians and five PAs (33% of total eligible providers) to participate in the semi-structured interviews. Four of the PAs and three of the Physicianss held leadership roles in HM at some point during the pre-or post-implementation phase of the care model. At the time of care model implementation, the interviewees had an average of five years of experience with the HM team.

During the interviews, participants discussed thirty-nine constructs of the CFIR framework. The three most frequently discussed constructs, and present in all ten interviews, were **work infrastructure, reflecting and evaluating, and innovation deliverers**. Additionally, all the APPs identified **teaming and culture** themes in their interviews. The theme of **access to knowledge** also was frequently discussed across provider types.

**Figure 1** demonstrates the most common themes by frequency discussed by provider type. **Figure 2** represents aggregate scores based on rapid analysis coding and consensus. This visual comparison is used to highlight the strength of impact as well as positive or negative influence of different themes by provider type.

**Figure 1:**
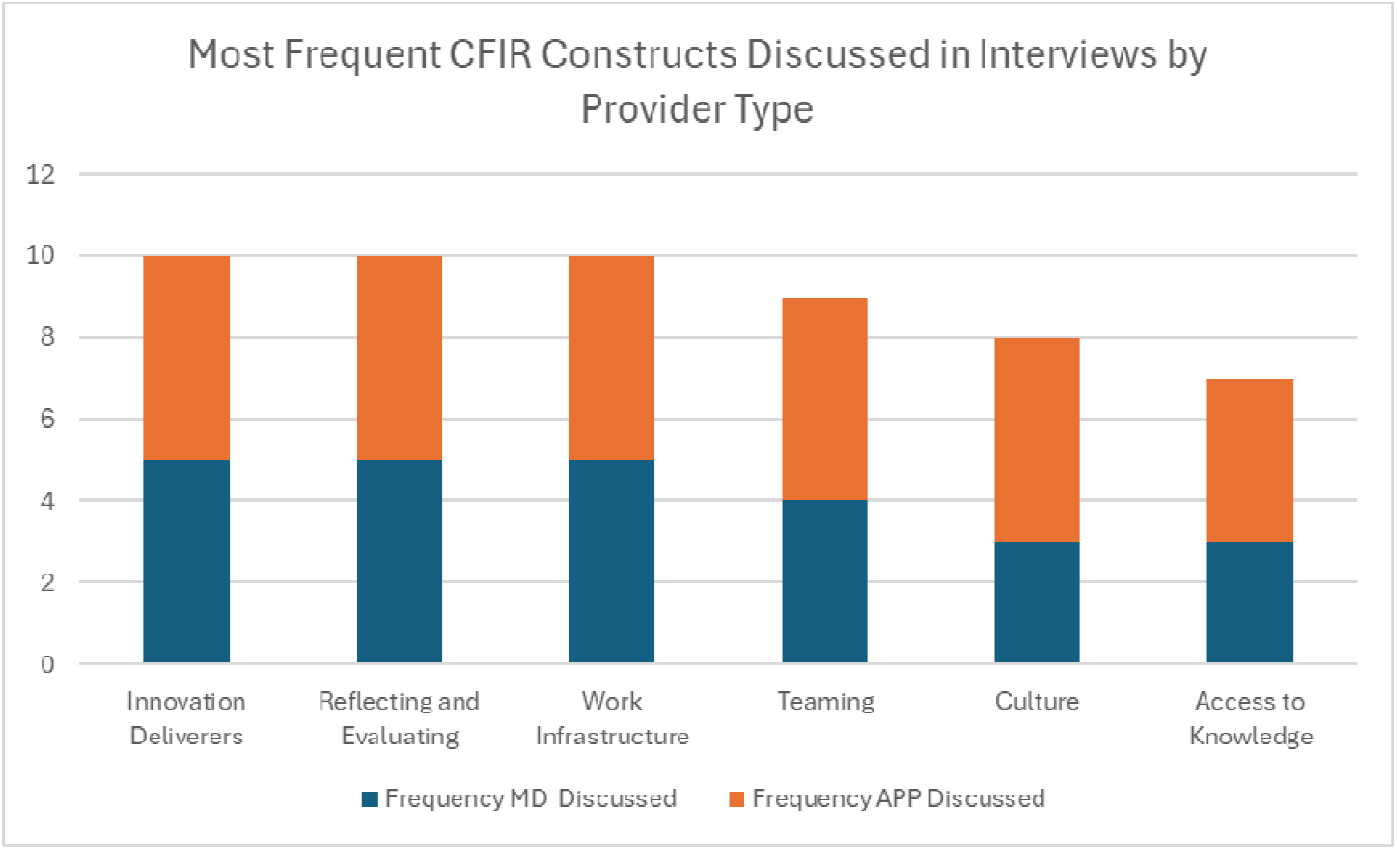
Frequency of themes discussed by provider type.

**Figure 2:**
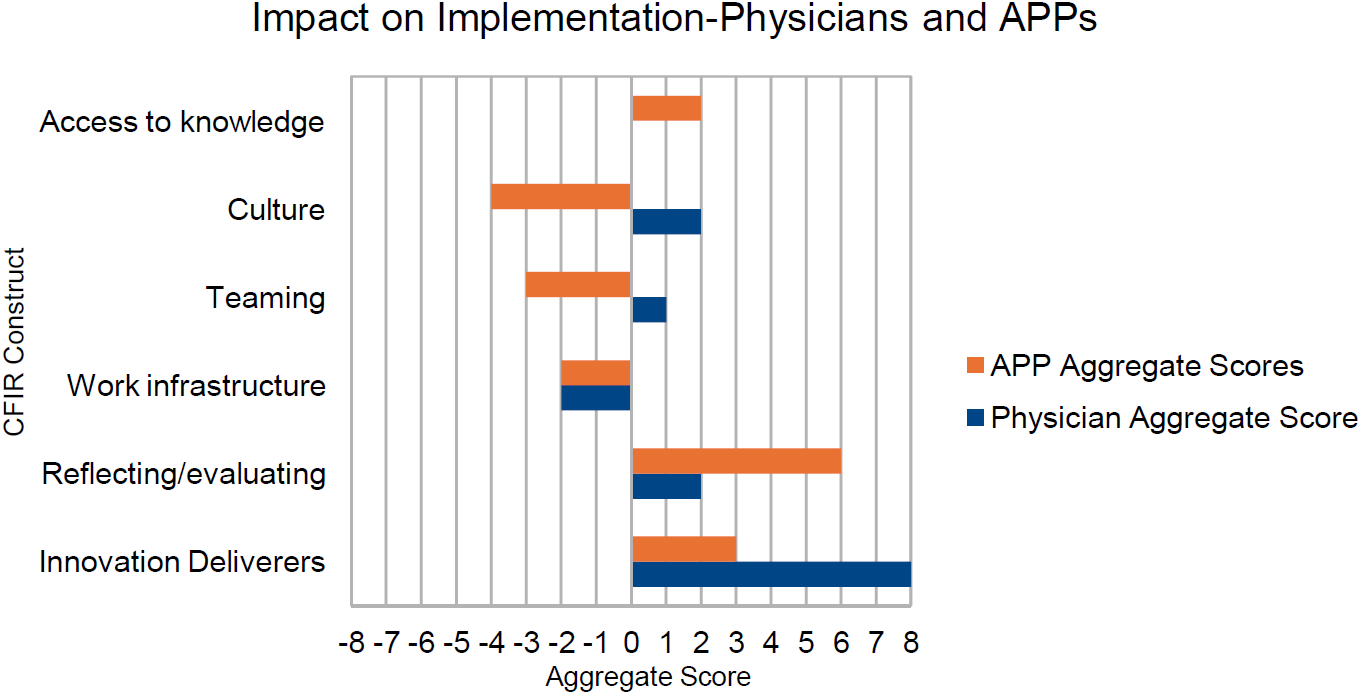
Aggregate score by provider type on most frequent CFIR Construct.

There were six primary themes identified. Two themes emerged as related to the strength of the care model implementation and shared in **Table 1** with exemplar quotes.

**Table 1:** Themes related to strength of care model implementation.

| Constructs (Domains) | Themes | Exemplar Quotes by Provider Type |
| --- | --- | --- |
| Innovation Deliverers (Individuals): | Experienced APPs delivered the new care model. | Regarding care model design, you need *“the right people on the bus before you start driving.”* -Physician<br>*“As an individual APP, I think the greatest strength is that I do get the opportunity to practice at the top of my license and really get to, you know, carry my own patients and that trust between myself and a physician as well as my patients and then other clinical staff is there and that’s very rewarding.”* - APP<br>*“The care model plays to the strength of the experienced APP (2-3 years great, 4-5 years better, >8 years best).”* -APP |
| Reflecting and Evaluating (Implementation Process): | Acceptance of the APP care model evolved over time. | *“If advising another team; talk through potential challenges; trust has to be built so physicians feel comfortable with safe APP care; build the program best suited to your location, culture, experience of APPs, workflow and patients seen.”* - Physician<br>*“Although reception of the model was slow, it made tremendous progress, 180-degree shift for both physicians and APPs.”* -APP<br>*“You have to ensure the right people are hired, invested in your mission, and accountable for the care model to succeed.”* -Physician |

### Theme 1: Experienced APPs delivered the new care model

Physicians remarked at the time of implementation, the APPs were experienced and they had confidence in their clinical acumen and supported their professional growth. Physicians also stated there was less duplicative work after the new care model was implemented, highlighting the efficiency of the new model. APPs stated they found reward in autonomy and accountability, which allowed physicians to focus their expertise on patients who needed it.

### Theme 2: Acceptance of the APP care model evolved over time

During initial implementation there was some skepticism by physicians of the ability for APPs to deliver care, over time there was overall acceptance by HM and other services of the expanded APP role. This was related to the physician and APP confidence in the care model.

In contrast, four themes emerged as opportunities for improvement of the care model implementation as highlighted in **Table 2** with exemplar quotes.

**Table 2:**
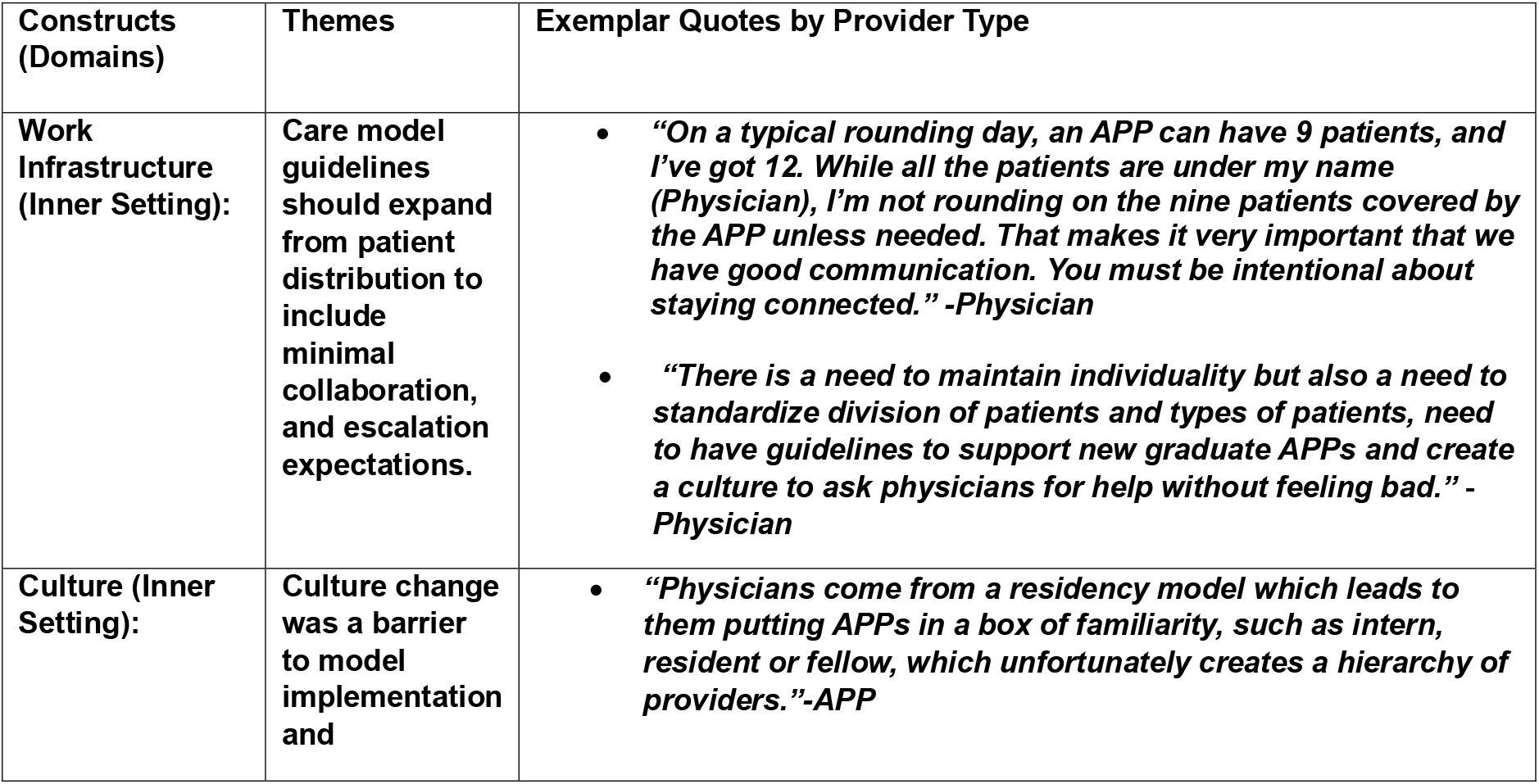
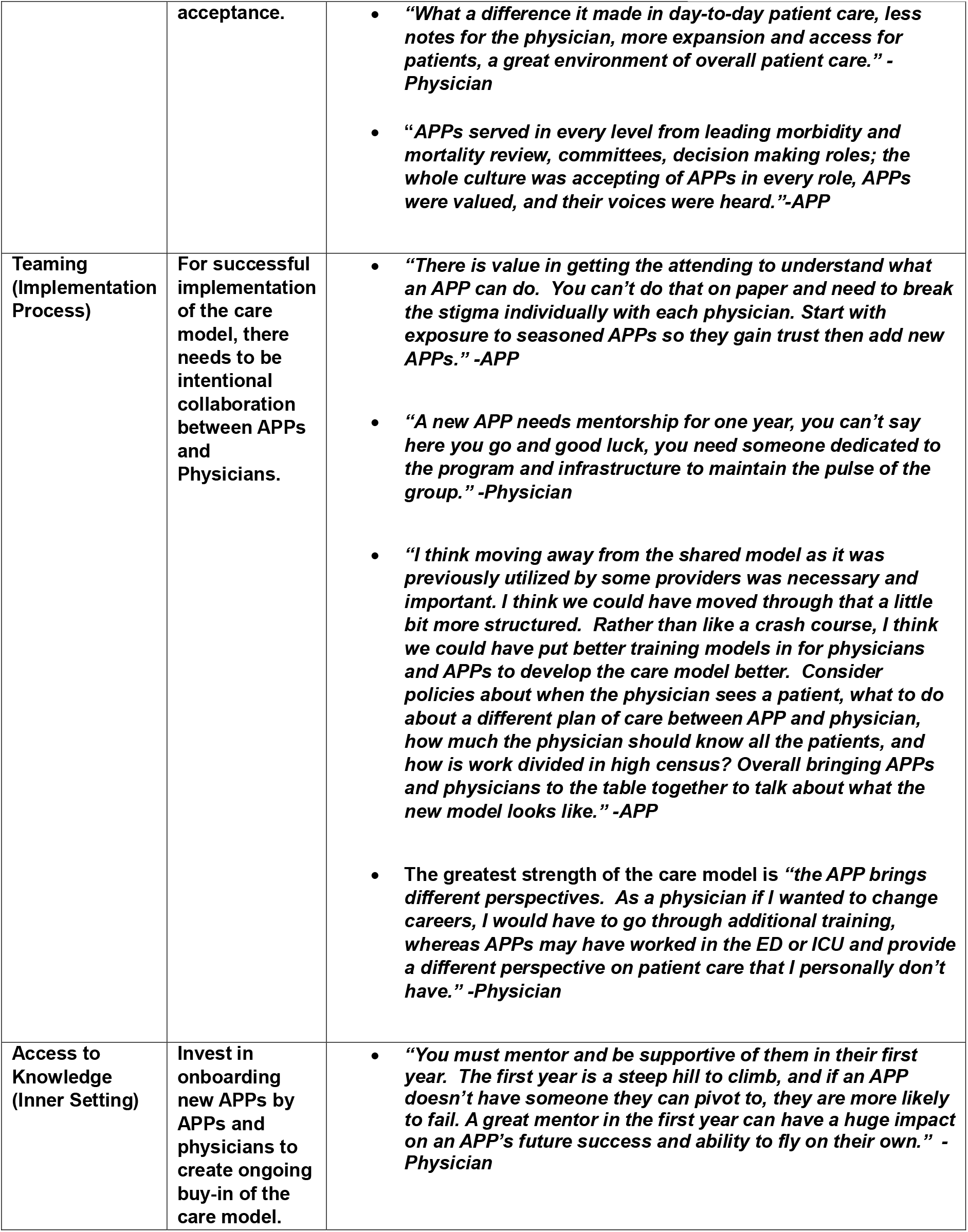
Themes related to opportunity for improvement of the care model implementation.

### Theme 3: Care model guidelines should expand from patient distribution to include minimal collaboration and escalation expectations

The care model was initially developed to meet increased patient demands from the pandemic. There were not enough physicians to meet the patients’ needs, necessitating optimizing care delivered by physicians and APPs. Initially, the only guideline for the care model was APPs were assigned a maximum of nine patients, and the physician took the rest of the patients to a maximum of 14. However, ultimately the physician was responsible for all service patients even if only in chart review and reliance on APP autonomy in the care model. In general, the APP decided which patients they would like to care for, without definitive standards of patient selection based on geography, acuity, or familiarity from past admissions. This inadvertently created dissatisfaction for some physicians who felt the distribution favored more difficult patients going to physicians. Additionally, it created friction when a physician questioned the unfair load of patients, without consideration of the hospital medicine service guidelines on APP management of 9 patients. Another opportunity identified was in the process of APP and physician collaboration, it varied by physician preference and led to more hands-off division of work. The APP suggested it would be best practice to discuss the list with the physician but preserve APP autonomy in driving which patients need seen by the physician based on complexity. The physician opinion was that it should be required to involve the physician for support if a patient is decompensating, even if the APP still leads the care. This ambiguity identified the need to define the process of organization of tasks and responsibilities between the APPs and physicians to preserve culture.

### Theme 4: Culture change was a barrier to model implementation and acceptance

Younger physicians who reported prior experience working with APPs adapted more easily compared to those who were new to working with APPs. Those who were skeptical of the care model had flexibility to do more than was required, such as seeing all patients and writing notes. This variance had the potential to erode trust in the model.

A strong APP leadership team provided structure and support for the care model. APPs had increased responsibility and growth, and the APP leader provided accountability to expectations and changes in role. The APP leaders demonstrated to the rest of the clinical team the APP potential in the clinical role as well as expanded opportunities for APPs leading committees and other quality improvement roles. When the model is thriving, it is very positive and beneficial to the work-life balance of physicians and APPs.

### Theme 5: For successful implementation of the care model, there needs to be intentional collaboration between APPs and Physicians

The APP care model was started in response to an external factor. The Covid-19 pandemic required dramatic changes due to increased patient volumes and acuity requiring the entire team to work to their top of scope. Participants remarked there was no significant onboarding to this new model, it just transitioned to an APP-led model of care. Inadvertently, physicians and APPs became more separated, and less teaming occurred in implementation. This included separate physician and APP meetings, perception of inequity of work division and responsibilities and barriers in communication. Another important missing piece was an opportunity for bi-directional stakeholder feedback.

### Theme 6: Invest in onboarding new APPs by APPs and Physicians to create ongoing buy-in of the care model

Throughout the interviews it was clear the care model worked well with experienced APPs with supportive physicians. As new members were added to the team, it became clear that new graduate APPs do not have the same level of experience, intuition and confidence. There is a need for investment in new APP onboarding with expanded medical knowledge and training tests, didactic lectures, and M&Ms to review patient cases. There should be shared responsibility of APP and physician peers for formal guidelines of ramp up to an autonomous role. By having an experienced APP and physician mentor, the APP felt more comfortable asking questions and grew in confidence, as well as the team benefited. Attrition of teams is a natural part of the cycle so supporting transition ensures stability of the care model.

## DISCUSSION

Both physicians and APPs who participated in the care model perceived it as successful in improving quality patient care and overall provider satisfaction. The model was built with experienced APPs which accelerated adoption of the new care model, however with robust standardized onboarding, patient care guidelines and multi-disciplinary mentorship, new APPs also have potential for success in the HM care model. The providers identified that care model implementation requires cultural change, which relies on trust building, communication and partnership between APPs and physicians to be successful. An APP Care Model cannot be built and left to evolve on its own, administrative leadership, APPs and physicians must intentionally collaborate to maximize success based on team feedback to reduce barriers to acceptance.

There were no limits placed on the APP role in this new model to focus on admissions, discharge planning or care of lower acuity patients, compared to other HM APP care models.^23^ These APPs served as primary HM care providers for a subset of HM service complex patients. Both physicians and APPs expressed job satisfaction with this model if teamwork was upheld; participants in our study reported lower satisfaction with the new care model if they felt isolated or siloed.

Our results are consistent with prior literature which found that APP utilization and deployment is usually preceded by external factors while failing to account for team dynamics and complexity.^7^ The HM APP-led care model that we studied also started due to a global pandemic and not planned iterative changes, which could account for negative impacts to teamwork in the model. The HM APP care model at our institution is evolving and incorporating IS principles such as stakeholder engagement, planned design and phased implementation throughout the evolution to improve the care model, provider satisfaction, and team collaboration. Other principles of change management, such as Kotter’s 8 Steps Process for Change could also be helpful for new services. ^24^ An important design element missing was the opportunity for formal feedback to allow for pivots in the care model, which eroded teaming and culture, especially from the APP perspective.

Ideas for future research include implementation of a hospital-based APP care model utilizing IS change management principles from the beginning. Evaluating the new APP care model would be based on measuring pre- and post-implementation team and individual provider productivity, quality care outcome measures such as re-admissions, length of stay, mortality, patient and provider satisfaction.

A potential limitation of the study was that it was conducted at a single AMC, which could have influenced cultural change and team dynamics. Only physicians and PAs participated in the qualitative interviews; the NP perspective was represented by a member of the study team who influenced the development throughout this study. The frontline NP voice should be represented in future studies. Seven of the providers interviewed reported holding a HM leadership role, which may have influenced their perspectives. Additionally, including more frontline provider perspectives would ensure a more thorough evaluation of the care model.

In today’s evolving healthcare landscape, it is imperative that all inpatient providers optimize the care and efficiency they provide. Expanding APP roles in inpatient care has the potential to increase efficiency while maintaining high quality care. The strengths and opportunities of care model implementation identified in this study provide a foundation for future care model design for HM teams. The same principles and concepts can be modified to match other specialty care model design to enhance perceptions of quality patient care and provider satisfaction.

## Data Availability

All data produced in the present work are contained in the manuscript

## ACKNOWLEDGEMENTS

A pre-press copy of this manuscript has been posted to the medRxiv.org pre-print server.

This manuscript was created as part of graduation requirements for the DMSc program of Wake Forest University School of Medicine Physician Assistant Studies.

This study would not have been possible without the HM physicians, physician assistants and nurse practitioners involved in the APP care model and the patients cared for by this team.

Additional gratitude is given to the colleagues in Wake Forest Clinical Translational Science Institute who gave consultation on qualitative research strategies, with special recognition to Dr Rachel Zimmer who provided mentorship.

## Notes

### Competing Interest Statement

The authors have declared no competing interest.

### Funding Statement

This study did not receive funding.

### Author Declarations

The Wake Forest University School of Medicine Institutional Review Board gave ethical approval for this study.

## REFERENCES

1) Wachter RM, Goldman L. The emerging role of “hospitalists” in the American health care system. N Engl J Med. 1996;335(7):514–517.

2) Džakula A, Lončarek K, Hass L, Vočanec D. Hospitalists: the missing link in complex patient care. Croat Med J. 2023;64(5):374–376.

3) Bryant SE. Filling the gaps: Preparing nurse practitioners for hospitalist practice. J Am Assoc Nurse Pract. 2018;30(1):4–9.

4) Zhao Y, Quadros W, Nagraj S, Wong G, English M, Leckcivilize A. Factors influencing the development, recruitment, integration, retention and career development of advanced practice providers in hospital health care teams: a scoping review. BMC Med. 2024;22(1):286. Published 2024 Jul 8.

5) https://www.nccpa.net/wp-content/uploads/2025/05/2024-Statistical-Profile-of-Board-Certified-PAs.pdf. Accessed February 4, 2026.

6) Reed, D, et al. The Pivotal Role of Nurse Practitioners as Hospitalists: Opportunities and Challenges. Nurse Leader, Volume 23, Issue 1, 44–47.

7) Westergaard S, Bowden K, Astik GJ, et al. Impact of billing reforms on academic hospitalist physician and advanced practice provider collaboration: A qualitative study. J Hosp Med. 2024;19(6):486–494.

8) McGrath BA, Jacobs ML, Watts RM, Callender BC. Developing and sustaining advanced practice provider services: A decade of lessons learned. J Hosp Med. 2022;17(12):1014–1020.

9) Keniston A, Patel V, McBeth L, Bowden K, Gallant A, Burden M. The impact of surge adaptations on hospitalist care teams during the COVID-19 pandemic utilizing a rapid qualitative analysis approach. Arch Public Health. 2022;80(1):57. Published 2022 Feb 17.

10) Nelson A, Fox J, Toth H, Stephany A. Ramping Up a Pediatric Hospital Medicine Advanced Practice Provider Team Rapidly. Hosp Top. 2021;99(1):44–47.

11) Lappé KL, Raaum SE, Ciarkowski CE, Reddy SP, Johnson SA. Impact of Hospitalist Team Structure on Patient-Reported Satisfaction with Physician Performance. J Gen Intern Med. 2020;35(9):2668–2674.

12) Johnson SA, Ciarkowski CE, Lappe KL, Kendrick DR, Smith A, Reddy SP. Comparison of Resident, Advanced Practice Clinician, and Hospitalist Teams in an Academic Medical Center: Association With Clinical Outcomes and Resource Utilization. J Hosp Med. 2020;15(12):709–715.

13) DeWolfe C, Birch S, Callen Washofsky A, Gardner C, McCarter R, Shah NH. Patient Outcomes in a Pediatric Hospital Medicine Service Staffed With Physicians and Advanced Practice Providers. Hosp Pediatr. 2019;9(2):121–128.

14) Gourley B, Akintade B, Appleby T, Bindon S, Idzik S. Developing Nurse Practitioners for Hospitalist Roles: Lessons Learned From an Academic Practice Partnership. J Dr Nurs Pract. Published online October 17, 2023.

15) Millwee S, Hall MAK. An Effective Model for Unit-Based Advanced Practice Provider/Physician Collaboration on a Complex Medicine Hospital Unit. J Nurs Adm. 2022;52(9):449–451.

16) Shannon EM, Cauley M, Vitale M, et al. Patterns of utilization and evaluation of advanced practice providers on academic hospital medicine teams: A national survey. J Hosp Med. 2022;17(3):186–191.

17) https://cancercontrol.cancer.gov/is/about#:~:text=Implementation%20science%20(IS)%20is%20the%20study%20of,activities%20to%20advance%20implementation%20research%20and%20practice. Accessed February 6, 2026.

18) Damschroder LJ, Reardon CM, Widerquist MAO, Lowery J. The updated Consolidated Framework for Implementation Research based on user feedback. Implement Sci. 2022;17(1):75. Published 2022 Oct 29.

19) Tong A, Sainsbury P, Craig J. Consolidated criteria for reporting qualitative research (COREQ): a 32-item checklist for interviews and focus groups. Int J Qual Health Care. 2007;19(6):349–357.

20) Reardon CM, Damschroder LJ, Ashcraft LE, et al. The Consolidated Framework for Implementation Research (CFIR) User Guide: a five-step guide for conducting implementation research using the framework. Implement Sci. 2025;20(1):39. Published 2025 Aug 16.

21) Hennink M, Kaiser BN. Sample sizes for saturation in qualitative research: A systematic review of empirical tests. Soc Sci Med. 2022;292:114523.

22) Nevedal AL, Reardon CM, Opra Widerquist MA, et al. Rapid versus traditional qualitative analysis using the Consolidated Framework for Implementation Research (CFIR). Implement Sci. 2021;16(1):67. Published 2021 Jul 2.

23) Dapaah-Afriyie K, Patel S, Beyer C. Advanced Practice Providers (APPs) in Hospital Medicine. Brown J Hosp Med. 2023;2(2):73047. Published 2023 Mar 14.

24) Campbell RJ. Change Management in Health Care. Health Care Manag (Frederick). 2020;39(2):50–65.

