## Appendix A for "Perceptions and Outcomes of a Hospital Medicine (HM) Advanced Practice Provider (APP)-Led Care Model: A Qualitative Study"

**Interview Guide**

***Thank you for taking the time to join us today. You have previously received a copy of the consent by email. Would you like me to read that to you? Do you have any remaining questions? Do you consent to participate?***

***I also want to confirm are you located in a place where we can have a private conversation without interruption.***

***As mentioned in the consent, I’m going to record our session. Your information will be kept in a secure file and will be recorded in a way that is anonymous and not linked to your name. Do I have your permission to record?***

***The primary investigator is present on the call for note taking but will not be interacting today.***

***Alright well let’s get started.***

***The interview is going to be broken into a few different sections so I may push us along to ensure we hit all areas.***

**Infrastructure and Innovation Source and Design**

1. Could you start by telling me about your current role in hospital medicine at the AFWFB Winston campus?
   a. Probe: When did you start in your role at Winston campus?
2. Reflecting on the period since 2020, what factors have influenced the development of today’s HM Physician and APP care model?
   a. Probes: What were the external pressures- events or pressures outside of the organization-that played a role (example if needed, workforce shortages, healthcare policy changes)?
   b. What were the internal factors- those arising from within the organization- that led to the adoption of this new model (example if needed, APP interest in expanded roles, efforts to improve workflow)?
   c. If pandemic mentioned, what aspects of the pandemic influenced this change?

**Implementation and Training**

3. Were you involved in designing or implementing the new model?
   a. If yes: What was your role in shaping the model?
4. Could you share how the transition was managed for APPs moving into the new care model? Probe: What training or structured support was offered to help APPs adjust?
5. How do physicians and APPs divide patient care responsibilities? Are there established rules or guidelines that determine which patients are assigned to an APP vs physician?
   a. If yes, could you describe these guidelines?
   b. If not, do you see opportunities to implement guidelines or suggest improvements?

**Culture**

6. How has collaboration between physicians and APPs changed over the time you have been involved?
7. Can you describe the typical daily workflow and the collaboration between physicians and APPs?
   a. Probe: Do you feel the current workflows are clearly structured and fully optimized?
   b. Is there a defined escalation pathway for APP-physician collaboration when dealing with a decompensating patient?
   c. If not: What factors prevent optimal collaborations, and what would you recommend improving?
8. What words would you use to describe the culture of hospital medicine as related to the APP-model of care?

**Reflection and Evaluation**

9. Have you noticed any outcomes, either positive or negative, that were unexpected with the implementation of this care model?
   a. Could you share specific examples?
10. How would you describe the overall culture and attitude within hospital medicine toward the APP model?

**Implementation and Future Planning**

11. What do you view as the greatest strength of this team-based care model?
12. What significant challenges or unexpected issues have you encountered with this model?
13. If you were advising another hospital service interested in adopting an APP-based model, what guidance would you provide?
14. Based on your experience, what lessons or important considerations should others keep in mind to avoid pitfalls?
15. Do you feel that this team-based care model enhances satisfaction for patients, APPs, and physicians? Could you share your insights or experiences?
16. Is there anything else we haven't covered that you feel is important to share?

*Thank you again for your time today. It is very much appreciated.*
